# A systematic review of (semi-)automatic quality control of T1-weighted MRI scans

**DOI:** 10.1101/2023.09.07.23295187

**Authors:** Janine Hendriks, Henk-Jan Mutsaerts, Richard Joules, Óscar Peña-Nogales, Paulo R. Rodrigues, Robin Wolz, George L. Burchell, Frederik Barkhof, Anouk Schrantee

## Abstract

Artifacts in magnetic resonance imaging (MRI) scans degrade image quality and thus negatively affect the outcome measures of clinical and research scanning. Considering the time-consuming and subjective nature of visual quality control (QC), multiple (semi-)automatic QC algorithms have been developed. This systematic review presents an overview of the available (semi-)automatic QC algorithms and software packages designed for raw, structural T1-weighted (T1w) MRI datasets. The objective of this review was to identify the differences among these algorithms in terms of their features of interest, performance, and benchmarks. We queried PubMed, EMBASE (Ovid), and Web of Science databases on the fifth of January 2023, and cross-checked reference lists of retrieved papers. Bias assessment was performed using PROBAST (Prediction model Risk Of Bias ASsessment Tool). A total of 18 distinct algorithms were identified, demonstrating significant variations in methods, features, datasets, and benchmarks. The algorithms were categorized into rule-based, classical machine learning-based, and deep learning-based approaches. Numerous unique features were defined, which can be roughly divided into features capturing entropy, contrast, and normative measures. Due to dataset-specific optimization, it is challenging to draw broad conclusions about comparative performance. Additionally, large variations exist in the used datasets and benchmarks, further hindering direct algorithm comparison. The findings emphasize the need for standardization and comparative studies for advancing QC in MR imaging. Efforts should focus on identifying a dataset-independent measure as well as algorithm-independent methods for assessing the relative performance of different approaches.

## 1. Introduction

Significant advances have been made in the realm of medical image analysis in the past few decades (Shaikh et al., 2020). Imaging biomarkers derived from advanced imaging techniques such as magnetic resonance imaging (MRI) data are used to characterize normal development (Oishi et al., 2013), disease (McEvoy and Brewer, 2010), and the effects of disease-modifying therapies (Paolillo et al., 1999). T1-weighted (T1w) MRI scans, for example, depict the anatomical arrangement of gray matter, white matter, and cerebrospinal fluid, providing valuable insights into the brain’s structural composition or pathology. However, before any image analysis workflow can be used, the quality of the MRI scans has to be ensured. Scan quality can be degraded by artifacts, which are unexpected/artificial image irregularities that are not related to anatomical or physiological abnormalities, but that can arise during the imaging process, such as blurring, ghosting, and aliasing. These artifacts can lead to low statistical power or erroneous conclusions.

Currently, quality of MRI data is often ensured through manual quality control (QC), which traditionally entails visual inspection of every individual scan of a dataset by an expert rater, from which those showing insufficient data quality are excluded. This manual QC is time consuming and prone to variability. Undesired variability may arise from inter- and intra-rater differences, such as training / experience and fatigue (Scheltens et al., 1995). Additional concerns are that subtle artifacts stemming from improper choice of acquisition parameters may be too subtle to be detected by the human eye (Gardner et al., 1995), or that differences in scanner vendor can introduce variability (Kruggel et al., 2010). These drawbacks of manual QC create great difficulty in defining objective exclusion criteria.

Furthermore, the acquisition of very large datasets across multiple scanning sites (Fantini et al., 2021; Sudlow et al., 2015; Van Essen et al., 2012) needed for clinical trials introduces additional concerns. Such large datasets make individual inspection of each image resource intensive, and add the possibility of between-site/rater variability. Therefore, there has been a great interest in the development of automated QC tools. Over time, several of these (semi-)automated QC algorithms have been created. Fully automated QC algorithms classify MRI scans without human involvement, while semi-automated QC algorithms require some level of human decision-making during the process, such as manually changing the threshold. Some of the algorithms focus on raw, unprocessed MRI scans (Alfaro-Almagro et al., 2018; Esteban et al., 2017) whereas others focus on their derivatives in the form of processed scans (e.g. segmentations) or statistics (e.g. regional volumetrics) (Keshavan et al., 2018; Klapwijk et al., 2019). However, despite their common goal of QC, these algorithms differ in their outcome parameters, and their application.

In light of the multitude of available tools, determining the most suitable choice can be challenging. Therefore, this review provides an overview of automated QC algorithms and software packages specifically designed for scrutinizing raw structural T1w MRI scans. We focus on whole brain, standard resolution T1-weighted MRI scans, typical of those ubiquitously employed in clinical trials and clinical practice. Our main objective is to identify the distinctions among these QC algorithms in terms of features of interest, performance, outcome metrics and type of data used as a benchmark.

## 2. Methods

Literature screening for this review was conducted according to Preferred Reporting Items for Systematic Reviews and Meta-Analyses (PRISMA) 2020 Guidelines (Liberati et al., 2009; Moher et al., 2010; Moher et al., 2015) and was registered with the Prospective Register of Systematic Reviews (PROSPERO) database under number CRD42023391301.

### 2.1. Data Sources and Searches

A systematic search was conducted to identify algorithms for (semi-)automatic quality control of structural MRI scans. Formal methods for literature search, selection, quality assessment, and synthesis were used, according to the PRISMA Guidelines (Liberati et al., 2009; Moher et al., 2010; Moher et al., 2015). The systematic search was performed in the databases: PubMed, Embase.com and Clarivate Analytics / Web of Science Core Collection. The timeframe within the databases was from inception to 5th January 2023 and the initial search was conducted by GLB and JH. The search included keywords and free text terms for (synonyms of) ’artificial intelligence’ or ’machine learning’ combined with (synonyms of) ’quality control’ combined with (synonyms of) ’ Magnetic Resonance Imaging ’ combined with (synonyms of) ’neuroscience’. A full overview of the search terms per database can be found in the supplementary information (Supplementary Tables 1 - 3). No limitations on date or language were applied in the search. Additionally, reference lists of included articles were manually screened to identify additional articles.

### 2.2. Article inclusion and exclusion criteria

Articles were included in this review if the following inclusion criteria were met: (1) involved the performance of (semi-)automatic quality control, (2) evaluated raw T1-weighted human brain MRI scans, and (3) published in English as original research in peer-reviewed journals or conference proceedings (conference abstracts and posters excluded). Articles involving QC of the output of preprocessing pipelines, as well as articles simply applying existing QC algorithms were excluded.

### 2.3. Data synthesis and analysis

To assess the eligibility of the selected articles, two independent reviewers (JH and AS) reviewed all abstracts from the database searches and retrieved full-text articles for further review. Any discrepancies were resolved through consensus. Finally, one reviewer (JH) read the retrieved articles for final article selection and quality assessment. The bibliographies of the retrieved full-text articles were manually searched for additional publications. For quality assessment, the PROBAST (Prediction model Risk Of Bias ASsessment Tool) (Wolff et al., 2019) method was chosen, being commonly used for assessing the risk of bias and applicability of prediction model studies. In this tool, risk of bias is defined to occur when shortcomings in the study design, conduct, or analysis lead to systematically distorted estimates of the model’s performance. Concerns regarding applicability of an article to the review question can arise when the population, predictors, or outcomes differ from those specified in the review question.

### 2.4. Data extraction

All full text articles that met the inclusion criteria were assigned into one of the following three categories: “Rule-based”, “Classical Machine learning” and “Deep learning”, based on the classification method used in the described algorithm. Rule-based algorithms establish thresholds for pre-defined quality features, while classical machine learning algorithms utilize a classifier to differentiate two groups based on an empirically established threshold from predefined quality features. Deep learning algorithms differ from those two groups as they do not rely on pre-defined quality features, and use a classifier to determine the groups. All articles were assessed by one rater and technical information and features of the algorithms including used datasets, age-range of included participants, artifact presence, benchmark, QC result, and performance measures were extracted. When possible, missing performance measures were calculated manually from data available in the articles.

## 3. Results

### 3.1. Search Results

The electronic search yielded 268 hits from PubMed, 496 from EMBASE, and 252 from Web of Science, amounting to a total of 1,016 hits (Figure 1). After removing duplicates, 605 unique articles were identified. After title and abstract review, 576 articles were excluded, and 29 were sought for retrieval of which one could not be retrieved. Of the 28 articles that underwent full text review, 12 were excluded from further quality assessment because of 1) assessing the quality of already processed (rather than raw) scans (n = 5), 2) relying on visual QC only (n = 3), 3) evaluating previously published algorithms (n = 2), 4) assessing quality of the file structure (n = 1) and 5) assessing quality of other MR sequences (n = 1). Cross-reference searching of the included articles resulted in the identification of eight more articles, of which six underwent full text review of which four were excluded from further quality assessment due to 1) assessing quality of other MR sequences (n = 2), 2) assessing quality of a file structure (n = 1) and 3) assessing quality of phantom images (n = 1). Ultimately, a total of 18 articles were included (Alfaro-Almagro et al., 2018; Bottani et al., 2022; Esteban et al., 2017; Fantini et al., 2021; Gedamu et al., 2008; Ikushima et al., 2022; Jang et al., 2018; Keshavan et al., 2019; Kim et al., 2019; Küstner et al., 2018; Mortamet et al., 2009; Osadebey et al., 2017a; Osadebey et al., 2017b; Osadebey et al., 2018; Pizarro et al., 2016; Sujit et al., 2019; White et al., 2018; Woodard and Carley-Spencer, 2006). Relevant information regarding dataset, benchmark, and performance measures is summarized in Table 1.

**Figure 1:**
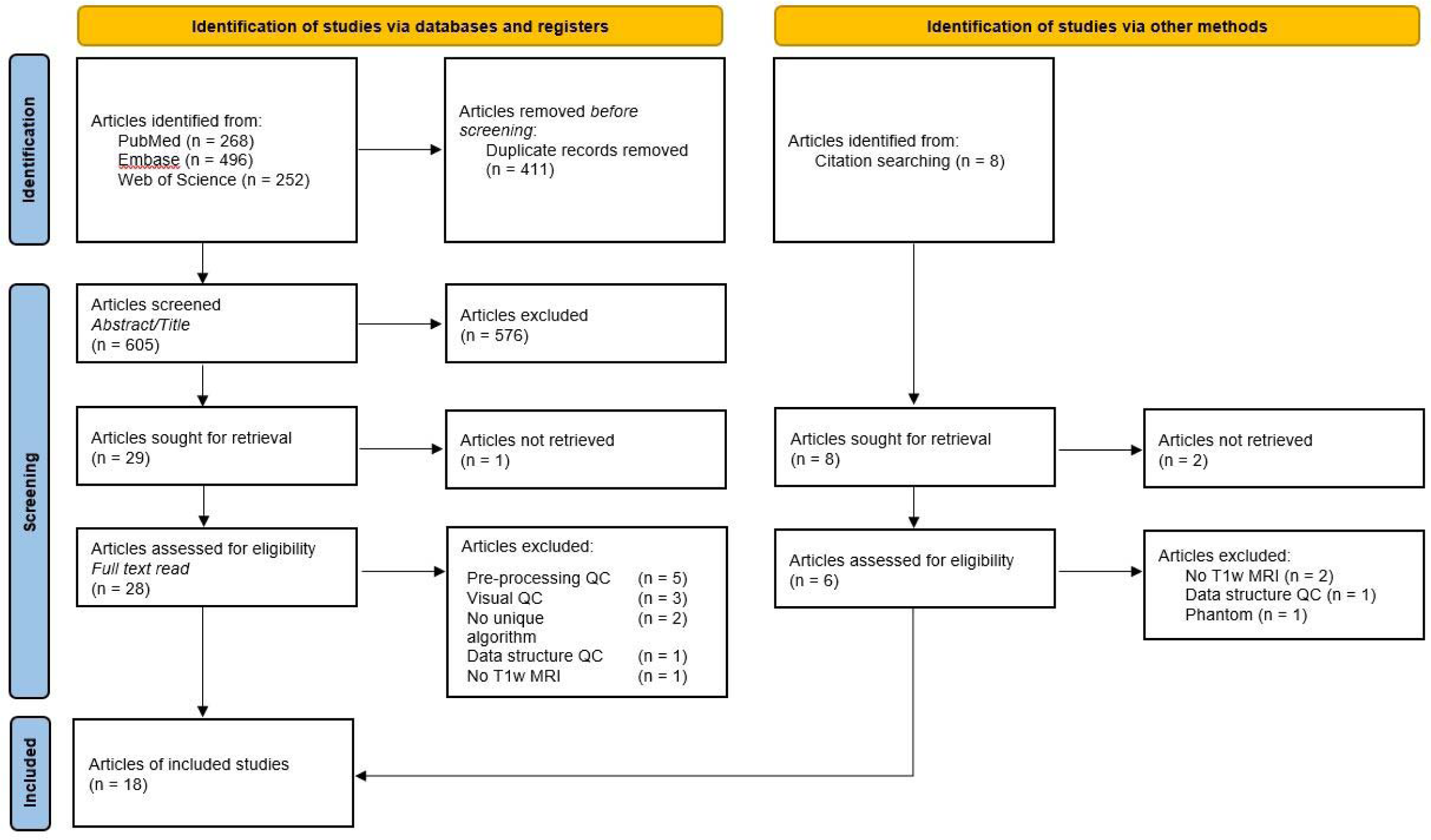
PRISMA 2020 flow diagram for new systematic reviews which included searches of databases, registers and other sources. *From:* Page MJ, McKenzie JE, Bossuyt PM, Boutron I, Hoffmann TC, Mulrow CD, et al. The PRISMA 2020 statement: an updated guideline for reporting systematic reviews. BMJ 2021;372:n71. doi: 10.1136 / bmj.n71. For more information, visit: http://www.prisma-statement.org/

**Table 1:** Data extraction.

| Article | Method<br>(# of features) | Dataset |  |  |  | benchmark | QC result | Performance measures |  |  |  |  |  |
| --- | --- | --- | --- | --- | --- | --- | --- | --- | --- | --- | --- | --- | --- |
|  |  | Name | N | Age-range (y) | Artifact presence (%) |  |  | Sensitivity / Specificity | Accuracy | PPV / NPV | BA | AUC | Pearson's R |
| (Gedamu et al., 2008) | RB (2) | NA | 40 | n.k. | NA | Visual | SNR Pass / Fail | NA | NA | NA | NA | NA | 0.98 |
|  |  | NA | 250 | n.k. | 52.4 | Visual | Ghosting yes / no | 93.2 / 92.3 | NA | NA | 92.8* | NA | NA |
|  |  | NA | 50 | n.k. | NA | Visual | Ghosting yes / no | 95.5 / 96.4 | 96.0* | 95.5* / 96.4* | 96.0* | NA | NA |
| (Ikushima et al., 2022) | RB (20) | NA | 100 | 25 - 38 | NA | Visual | Ranking | NA | NA | NA | NA | NA | 0.92 |
| (Jang et al., 2018) | RB (20) | ADNI + ABIDE | 50 | 55 - 90 + 5 - 64 | NA | Visual | Ranking | NA | NA | NA | NA | NA | 0.93 - 0.95 |
| (Kim et al., 2019) | RB (7) | TRACK-TBI | 1569 | 18 - 39 | 1.59 | Visual | Pass / Fail | 85.0 / 87.0 | 89 | NA | 86.0* | 0.93 | NA |
| (Mortamet et al., 2009) | RB (2) | ADNI + ABIDE | 749 | 55 - 90 | 7.21 | Visual | Pass / Fail | 87.2 / 85.2 | NA | NA | 86.2* | 0.93 | NA |
| (Osadebey et al., 2017b) | RB (3) | ADNI + NeuroRX | n.k. | 55 - 90 + n.k. | NA | Simulated | Ranking | NA | NA | NA | NA | NA | NA |
| (Osadebey et al., 2018) | RB (2) | ADNI + NeuroRX | n.k. | 55 - 90 + n.k. | NA | Simulated | Ranking | NA | NA | NA | NA | NA | 0.73 |
| (Osadebey et al., 2017a) | RB (5) | NeuroRX | n.k. | n.k. | NA | Simulated | Ranking | NA | NA | NA | NA | NA | NA |
| (White et al., 2018) | RB (3) | Generation R Wave I | 1070 | 6 - 11 | 4.77 | Visual | Pass / Fail | NA | NA | NA | NA | 0.95 | NA |
|  |  | Generation R Wave II | 4339 | 8 - 12 | 8.78 | Visual | Pass / Fail | NA | NA | NA | NA | 0.95 | NA |
|  |  | NHGRI | 442 | 5 - 78 | 5.2 | Visual | Pass / Fail | NA | NA | NA | NA | 0.88 | NA |
|  |  | GUSTO | 811 | 4 - 55 | 54.62 | Visual | Pass / Fail | NA | NA | NA | NA | 0.82 | NA |
| (Woodard and Carley-Spencer, 2006) | RB (259) | MNI | 143 | n.k. | NA | Simulated | Pass / Fail | NA | NA | NA | NA | NA | NA |
| (Alfaro-Almagro et al., 2018) | ML (190) | UK Biobank | 5816 | 40 - 60 | 1.77 | Visual | Pass / Fail | 91.3 / 84.0 | 84.2* | 9.3 / 99.8 | 87.6 | NA | NA |
| (Esteban et al., 2017) | ML (64) | ABIDE | 1102 | 5 - 64 | 30.58 | Visual | Pass / Fail | NA | 76.2 | NA | NA | 0.73 | NA |
|  |  | DS030 | 265 | 21 - 55 | 28.3 | Visual | Pass / Fail | 76.9* / 67.7* | 75.8 | 94.7* / 28.0* | 72.3 | 0.71 | NA |
| (Pizarro et al., | ML (6) | CBDB | 1457 | 18 - 60 | 13.04 | Visual | Pass / Fail | 70.1 / 88.2 | 79 | NA | 79.2* | NA | NA |
| 2016) |  | Sibling Study |  |  |  |  |  |  |  |  |  |  |  |
| (Bottani et al., 2022) | DL | NA | 3770 | 18 - 96 | 43.47 | Visual | Pass / Fail | 79.9 / 87.1 | 84.2* | 81.9 / 85.8 | 83.5 | 83.5 | NA |
|  |  |  | 2182 |  | 40.01 | Visual | Medium / good | 77.4 / 65.9 | 73.9* | 83.2 / 57.8 | 71.7 | 71.7 | NA |
| (Fantini et al., 2021) | DL | NA | 68 | 21-61 | 50 | Visual | Pass / Fail | NA | 90.4 | NA | NA | NA | NA |
|  |  | NA | 93 | 8 - 66 | 40.86 | Visual | Pass / Fail | 97.4 / 85.5* | 90.3 | 82.2 / 97.9* | 91.4* | NA | NA |
|  |  | NA | 22 | 19 - 65 | 9.09 | Visual | Pass / Fail | 100 / 60.0* | 63.6 | 20.0 / 100* | 80.0* | NA | NA |
|  |  | NA | 20 | 22 - 85 | 30 | Visual | Pass / Fail | 100 / 71.4* | 75 | 33.3 / 100* | 85.7* | NA | NA |
| (Keshavan et al., 2019) | DL | HBN | 200 | 5 - 18 | 50 | Visual | Pass / Fail | NA | NA | NA | NA | 0.99 | NA |
| (Kustner et al., 2018) | DL | NA | 32 | n.k. | 50 | Visual | Motion yes / no | NA | 92 | NA | NA | NA | NA |
| (Sujit et al., 2019) | DL | ABIDE | 1064 | 5 - 64 | 12.41 | Visual | Pass / Fail | 77.0 / 85.0 | 84 | 42.0 / 96.0 | 91.0* | 0.9 | NA |

n.k. = not known, NA = not available, RB = rule-based, ML = machine-learning based, DL = deep learning based, PPV = positive predictive value, NPV = negative predictive value, BA = balanced accuracy, AUC = area under the curve, \* = manually calculated performance measure

### 3.2. Risk of bias assessment

All included articles showed a low concern regarding applicability of the algorithms (Table 2). For 16 articles, the outcome measures of the articles showed a high risk of bias, since the benchmarks are mostly based on a visual QC (Table 1), which can lead to distorted or flawed assessment in the benchmarks. Examples of factors introducing risk of bias for a visual QC approach are an unclear protocol (Alfaro-Almagro et al., 2018; Jang et al., 2018; Mortamet et al., 2009), undisclosed number of raters (Alfaro-Almagro et al., 2018; Gedamu et al., 2008; Mortamet et al., 2009) or QC based solely on the assessment of one rater (Fantini et al., 2021; Kim et al., 2019; Pizarro et al., 2016). Moreover, the analysis of various algorithms are at high risk of bias, due to lack of validation (Gedamu et al., 2008; Ikushima et al., 2022; Jang et al., 2018; Mortamet et al., 2009; Osadebey et al., 2017a; Osadebey et al., 2017b; Osadebey et al., 2018; Woodard and Carley-Spencer, 2006) or a low number of participants (Fantini et al., 2021; Ikushima et al., 2022; Jang et al., 2018; Küstner et al., 2018).

**Table 2:** Risk of Bias Assessment.

| Article | ROB |  |  | Applicability |  | Overall |  |
| --- | --- | --- | --- | --- | --- | --- | --- |
|  | Predictors | Outcome | Analysis | Predictors | Outcome | ROB | Applicability |
| (Gedamu et al., 2008) | + | ? | - | + | + | - | + |
| (Ikushima et al., 2022) | + | + | - | + | + | - | + |
| (Jang et al., 2018) | + | - | - | + | + | - | + |
| (Kim et al., 2019) | + | - | + | + | + | - | + |
| (Mortamet et al., 2009) | + | - | - | + | + | - | + |
| (Osadebey et al., 2017b) | + | NA | - | + | NA | - | + |
| (Osadebey et al., 2018) | + | - | - | + | + | - | + |
| (Osadebey et al., 2017a) | + | NA | - | + | NA | - | + |
| (White et al., 2018) | - | - | + | + | + | - | + |
| (Woodard and Carley-Spencer, 2006) | + | NA | - | + | NA | - | + |
| (Alfaro-Almagro et al., 2018) | + | - | - | + | + | - | + |
| (Esteban et al., 2017) | + | - | + | + | + | - | + |
| (Pizarro et al., 2016) | + | + | + | + | + | + | + |
| (Bottani et al., 2022) | NA | + | + | NA | + | + | + |
| (Fantini et al., 2021) | NA | - | - | NA | + | - | + |
| (Keshavan et al., 2019) | NA | - | + | NA | + | - | + |
| (Küstner et al., 2018) | NA | - | - | NA | + | - | + |
| (Sujit et al., 2019) | NA | - | + | NA | + | - | + |

PROBAST = Prediction model Risk Of Bias ASsessment Tool; ROB = risk of bias; NA = not applicable. Domain 1: Participants was not relevant and hence disregarded. Domain 2 was disregarded for the algorithms utilizing a deep learning approach, since the predictors are not known a-priori in these cases.
\* + (green) indicates low ROB / low concern regarding applicability; - (red) indicates high risk of bias / high concern regarding applicability; ? (yellow) indicates unclear risk of bias / unclear concern regarding applicability

### 3.3. Rule-based QC

A total of ten articles were found that utilized a rule-based approach of one or more quality features to evaluate the quality of structural MRI scans (Gedamu et al., 2008; Ikushima et al., 2022; Jang et al., 2018; Kim et al., 2019; Mortamet et al., 2009; Osadebey et al., 2017a; Osadebey et al., 2017b; Osadebey et al., 2018; White et al., 2018; Woodard and Carley-Spencer, 2006). Eight articles categorize MRI scans into pass and fail groups (Ikushima et al., 2022; Jang et al., 2018; Kim et al., 2019; Mortamet et al., 2009; Osadebey et al., 2017a; Osadebey et al., 2017b; White et al., 2018; Woodard and Carley-Spencer, 2006), one assessed whether blurring is present (Osadebey et al., 2018), and one assessed SNR and ghosting without an overarching score (Gedamu et al., 2008). Five articles (Gedamu et al., 2008; Kim et al., 2019; Mortamet et al., 2009; White et al., 2018; Woodard and Carley-Spencer, 2006) evaluated quality features based on the background of the image, arguing that most of the artifactual signal intensities propagate over the image and into the background, which typically corresponds to 40% of the total volume of a structural MRI scan. The other articles (Ikushima et al., 2022; Jang et al., 2018; Osadebey et al., 2017a; Osadebey et al., 2017b; Osadebey et al., 2018) use foreground-based quality features, arguing that relying solely on the background may not provide a reliable measure of the overall image quality.

#### 3.3.1. Rule-based QC using background

One article used quality features stemming from previous studies trying to assess distortion in scans caused by image compression (Woodard and Carley-Spencer, 2006). In order to assess which of these quality features are most applicable for QC of MRI scans, they applied a large set of quality features, classified into seven feature families, to artificially distorted MRI scans (N = 143). They found that quality features based on Natural Scene Statistics were the most effective in distinguishing between artificially distorted and undistorted MRI scans.

Subsequent studies investigated features specific for MR artifacts extracted from the background, such as noise and ghosting artifacts in scans (N = 250) (Gedamu et al., 2008). When these features were compared with white matter SNR and visually assessed ringing artifacts (undisclosed number of raters), very high sensitivity and specificity are found. They determined a QC threshold based on the highest agreement between the features and manual assessment. For the feature assessing ghosting, a validation test was performed, which resulted in a specificity and sensitivity similar to the training set.

Subsequent work (Mortamet et al., 2009), put forth the argument that SNR measures may not necessarily be sensitive to subject-related artifacts. Instead, they suggest that these artifacts lead to a corrupted noise distribution that can be evaluated using two specific quality features. The first feature assesses the effects of clustered artifacts in the background and the second feature evaluates both clustered and subtle effects of artifacts in the background (N = 749; undisclosed number of raters). White et al. (White et al., 2018) continued on this work, by calculating the integral of the voxel intensities as vectors radiating away from the head (N = 6662; 1 or 2 raters). This feature was compared with two other new features, which capture the frequency characteristics of the noise rippling away from the edge of the head and utilize properties of the line spread function along the edge of the head. In both these studies, visual quality assessment led to a binary pass or fail score, which was either used to determine the cut of values for the quality features (Mortamet et al., 2009) or to assess the performance by determining the area under the curve (AUC) (White et al., 2018). The feature utilizing the line spread function along the edge of the head was reported to perform best (White et al., 2018).

Finally, the LONI QC System (Kim et al., 2019) is a publicly available QC algorithm for structural T1w scans based on seven features, namely SNR, signal variance-to-noise variance ratio (SVNR), contrast-to-noise ratio (CNR), contrast of variance-to-noise ratio (CVNR), brain tissue contrast-to-tissue intensity variation (TCTV), full-width-at-half-maximum (FWHM), and center of mass (CoM). In this algorithm, the image background is used to represent noise. To assess the manual binary classification (one rater) accuracy of each QC feature, the values per feature was changed to a *z*-score, and a threshold was determined based on agreement with visual QC.

#### 3.3.2. Rule-based QC using foreground

Jang et al. (Jang et al., 2018) argue that not all MRI scans allow for capturing background noise, e.g., when the background area is insufficient to allow a robust analysis. They introduced the Quality Evaluation using MultiDirectional filters for MRI (QEMDIM) algorithm, which uses multidirectional filters to capture quality features. Each image is divided into 16 patches with 20 quality features, which were averaged over the patches. Image quality is determined by calculating the absolute difference between the averaged quality features of the test image and those of a benchmark of undistorted images, using the agreement with visual scores as threshold. Others (Ikushima et al., 2022) later modified QEMDIM such that it does not only provide the absolute quality difference but also if the assessed scan has a higher or lower quality than the benchmark. Additionally, calculation efficiency was improved by omitting patch division and feature averaging. Then they revalidated the modified QEMDIM score with a visual quality score.

Osadebey et al. (Osadebey et al., 2017a; Osadebey et al., 2017b; Osadebey et al., 2018) developed three algorithms to assess MRI scan quality, using foreground features. One algorithm (Osadebey et al., 2017a) calculates a total quality score as a weighted sum of noise, lightness, contrast, sharpness, and texture details. In a second algorithm (Osadebey et al., 2017b) three geo-spatial local entropy features are being extracted from all the slices of a MRI scans. In both these algorithms, it was shown that undistorted images have higher quality scores than artificially degraded images. In a more recent study (Osadebey et al., 2018), the authors used an average of a sharpness and a contrast quality feature; which also performed good in comparison to a visual rating scale.

### 3.4. Classical Machine Learning

Three studies applied classical machine learning approaches (Alfaro-Almagro et al., 2018; Esteban et al., 2017; Pizarro et al., 2016), which all classify MRI scans into either pass or fail by training them against visual QC results. Quality features based on the background and the foreground of the image have been used.

Pizarro et al. (Pizarro et al., 2016) investigated multi-dimensional, non-linear classification to overcome limitations of univariate approaches, including the need for multiple quality features to characterize artifacts from different sources, since a single quality feature has limited ability to capture details of artifacts in small local regions and cannot capture sufficient information on artefact type and location. Six different features were extracted; three volumetric features (related to contrast, intensity and tissue class) and three artifact-specific features (related to eye movement, ringing and aliasing). The MRI scans (N = 1457) are also visually assessed, and classified in either pass or fail by five to nine raters. The features and the visual assessment were fed to a supervised classification algorithm based on a support vector machine (SVM), which was trained with a 10-fold cross validation.

The UK biobank developed a QC algorithm to assess the quality of their own dataset only (Alfaro-Almagro et al., 2018). A classical machine learning approach was proposed to automatically identify problematic scans based on 190 image derived features. These features are derived from the raw images as well as after preprocessing. A Weka machine learning toolbox was used, with three separate classifier’s outputs fused together, and a voting system combining the a posteriori probabilities of the three classifiers was used for the fusion. To train this QC algorithm, the quality of the first release of the Biobank MRI scans (N = 5816) was assessed manually (amount of raters unknown). For training a stratified 10-fold cross validation was used. To test the algorithm, the second release of the Biobank was used, which was not manually labeled, and therefore no performance measures could be derived.

Esteban et al. (Esteban et al., 2017) developed MRIQC, a publicly available algorithm which extracts 64 image quality features, and fits a binary classifier. The features are based on background evaluation of the raw MRI scan only. A supervised classification framework was used, composed of a random forest classifier (RFC), with a Leave-one-Site-out splitting for cross-validation, where a whole site is left out as a test set at each cross-validation fold. The quality of all MR volumes (N = 1102) was first manually assessed by two raters, and they were given a label of ‘exclude’, ‘doubtful’ or ‘accept’. For the binary classifier, pass consisted of the scans with an ‘accept’ and ‘doubtful’ label, whereas fail was composed of all manually ‘excluded’ MRI scans.

### 3.5. Deep learning

Five studies utilized a deep learning approach (Bottani et al., 2022; Fantini et al., 2021; Keshavan et al., 2019; Küstner et al., 2018; Sujit et al., 2019), of which three classify the MRI scans as either pass or fail (Bottani et al., 2022; Keshavan et al., 2019; Sujit et al., 2019) and two (Fantini et al., 2021; Küstner et al., 2018) assess whether or not motion is present in the assessed scans.

The first article investigating the feasibility of automated detection and assessment of motion artifacts in MRI scans with a convolutional neural network (CNN) was published by Küstner and colleagues (Küstner et al., 2018). The input of the CNN was a patched image; it was also investigated which patch size was the best. For training and validation, MRI scans from 16 healthy volunteers were used. Each volunteer was scanned twice, and during the first acquisitions, the volunteers were instructed to hold their head still. During the second acquisition, volunteers were instructed to deliberately tilt their head side-to-side. For training and evaluation, a leave-one-out cross validation approach was used. This CNN was able to localize and detect motion artifacts. Furthermore, it was shown that patch-based accuracy of detecting motion artifacts declined with decreasing patch-size.

Fantini et al. (Fantini et al., 2021) continued on the work of Küstner (Küstner et al., 2018), by including scans with more complex motion distortions (N = 203). Furthermore, they also investigated the performance of four different networks, namely Xception, InceptionV3, ResNet50 and Inception-Resnet, and applied transfer learning by pretraining the networks on the Imagenet dataset. One model was trained for each standard MR orientation (axial, coronal and sagittal axes) on patches, and an artificial neural network was used to combine the outputs of the different networks to one output value. Also a depth search was performed, attaching the binary classifier on distinct blocks output and the best architecture depth was selected from the block layer that reported the best accuracy. A 3-fold cross-validation approach was used for the training of all architectures.

Sujit et al. (Sujit et al., 2019) aimed to develop an algorithm that would evaluate the image quality of 3D T1w MRI scans using data from a large multicenter database (N = 1064) which were classified by two raters. Their deep learning network was inspired by the VGG16 network, and one model was trained for each standard MR orientation (axial, coronal and sagittal axes), and the output layer provided a slice-wise quality score. A second network consisting of a fully connected layer and an output layer was used to combine all the slice scores into one “volumetric” quality score. It was shown that this model provided good accuracy for classifying brain MRI scan quality.

Bottani and colleagues (Bottani et al., 2022) developed an algorithm for automatic QC of MRI scans in a large clinical data warehouse (N = 3770) which were rated by two raters. They aimed to discard scans which are not proper T1w brain MRI, identify scans with gadolinium and recognise scans of bad, medium, and good quality. For the purpose of this paper, we will focus on the last two aims. MRI scans were classified into 3 tiers, i.e. good quality, medium quality and bad quality. For the classification between bad vs medium / good and medium vs good two separate networks were trained. It was trained using the cross entropy loss, which was weighted according to the proportion of scans per class for each task.

Another approach was used by Keshavan et al. (Keshavan et al., 2019) in which citizen scientists were used to visually QC MRI scans in order to acquire a large labeled dataset (N = 200) which in turn can be used to train deep learning networks for automatic QC. A VGG16 network was pretrained on the Imagenet database, and further training and testing was done with the scoring of both the experts and the citizen scientists. Performance was reported by comparing the outcomes of this network and amplified training dataset with the outcomes of MRIQC (Esteban et al., 2017).

## 4. Discussion

In this systematic review we identified 18 unique algorithms used for quality control of structural T1w scans, of which 10 use a rule-based approach, three use a classical machine learning approach and five use a deep learning approach. The results of our systematic review revealed three key findings. Firstly, we identified a wide array of features incorporated within these algorithms, and even though there is little consistency across algorithms in terms of features, most of them utilize at least one feature assessing the entropy of the image. Secondly, the lack of consistent metrics and evaluation criteria hindered direct comparisons and highlighted the importance of establishing standardized performance measures within the field. Lastly, we observed significant variability in the selection of benchmarks employed during the development of QC algorithms across different approaches.

### 4.1. Features

By design, both rule-based and machine-learning algorithms use pre-defined features aimed at capturing image properties. A great number of features have been proposed, but only signal-to-noise ratio (SNR) and contrast-to-noise ratio (CNR) have been used in three or more algorithms. The features can roughly be categorized into three categories: entropy features, image contrast features, and normative features. Initially studies focussed on entropy measures (Gedamu et al., 2008; Woodard and Carley-Spencer, 2006) capturing the randomness in an image, and therefore the majority of the features belong to this category because other studies build upon those. Features in more recent studies categorize best as normative measures, which capture deviations of the image compared to the mean. Features extracted from the background of an image can be classified into either the image contrast or entropy category. These categories are commonly utilized in studies evaluating noise or blur, which tend to distribute evenly throughout the image, thus including the background. Features extracted from the foreground generally belong to the normative category, with the purpose of identifying deviations from the usual patterns. Features related to normative features capture a broader range of artifacts than the other two categories. All algorithms combine features from different categories to detect a wide range of artifacts, except those using the QEMDIM algorithm (Ikushima et al., 2022; Jang et al., 2018). In that case the authors argue that a single feature, reflecting the distance from a benchmark, captures enough information to catch multiple sources for image degradation.

It should be noted that a minimal processing workflow is generally utilized to extract the different features, typically developed based on MRI scan of healthy individuals without significant artifacts. As a result, the presence of artifacts and clinical deviations can potentially impact the performance of the workflow and thus the extracted features. In cases where a scan fails the processing workflow, it could be attributed to the presence of artifacts or clinical deviations such as the presence of a tumor. This creates a circularity in feature extraction, where features are used to quantify artifact presence, yet the extraction of features is influenced by the presence of irregularities.

Deep learning algorithms do not use predefined features, and by design the factors that contribute to the classification by these networks are not always apparent, which limits interpretability. In the included articles, different architectures of CNNs are being used. Both traditional sequential network (Keshavan et al., 2019; Sujit et al., 2019) and network-in-network architectures are being used for QC (Fantini et al., 2021). Fantini et al. (Fantini et al., 2021) showed that the different architectures lead to different results, but only compared network-in-network architectures with each other. The different architectures were trained on the same dataset, and thus a fair comparison on performance could be made. Ultimately, deep learning algorithms can only be compared with each other, or other types of algorithms, based on the final performance measures.

### 4.2. Performance

One consistent finding is that the use of dataset-specific optimization creates a circular process that inflates performance measures. During training of the machine-learning and deep learning algorithms, or when setting the thresholds for the rule-based algorithms, an iterative approach is used in all studies that repeatedly refers back to the dataset for fine-tuning. This suggested that the performance of the algorithms was artificially inflated on that particular dataset. Consequently, comparing the performance of two or more algorithms becomes challenging, as the performance measures only reflect performance on specific datasets. The limitations of dataset-specific optimization become apparent when independent validation is conducted, causing the accuracy to decline with approximately 11%. Noteworthy, no independent validation of the thresholds in rule-based algorithms was performed, except for the ghosting threshold in (Gedamu et al., 2008). Independent validation is performed in a in one classical machine-learning algorithm (Esteban et al., 2017) and three deep learning algorithms (Bottani et al., 2022; Fantini et al., 2021; Sujit et al., 2019). As can be seen in Table 1, various performance measures have been used. Often accuracy was chosen to evaluate the model, since it coincides well with the general aim of developing a QC algorithm, i.e. to predict the class of unseen MRI scan accurately. However, it might not be the best performance measure in all cases, as it conveys well only when all classes (pass or fail) have similar prevalence in the data. However, this is not the case, as the majority of the studies utilizing visual QC have less than 40% of the used dataset categorized as fail. Additional measures like the sensitivity, specificity, PPV and NPV are therefore needed to provide a more complete overview of the performance.

### 4.3. Datasets

The specific characteristics of individual datasets introduce significant variations and pose challenges when comparing algorithms. The majority of the algorithms are trained on datasets incorporating participants with pathologies, especially Alzheimer’s disease (ADNI) and autism spectrum disorder (ABIDE), while some contain solely healthy individuals. The choice to include scans where pathologies might be present seems to be driven by the fact that those types of datasets are relatively big and publicly available. The studies including their own datasets often include solely healthy individuals, and are notably smaller. There is a risk that algorithms trained on healthy datasets may misclassify pathologies as artifacts (Krupa and Bekiesinska-Figatowska, 2015), although they may exhibit higher sensitivity to small artifacts. Variability in datasets is also found in the age distribution of the included participants. For two algorithms the age distribution is not reported (Gedamu et al., 2008; Osadebey et al., 2017b), but in the majority of utilized datasets adults (age range 18 - 65) were included. However, four datasets included children (age < 18) only, four included adults and elderly (age 18+), and two datasets a combination of children and adults (age < 65). It is shown that there are differences in cortical gray matter as a function of age between children and adults (Jernigan and Tallal, 1990), as well as that the cortical thickness shows regional and temporal specificity with development (Sowell et al., 2003). Differences are also found between adults and elderly, like atrophy or the expansion of the ventricular system (Fjell and Walhovd, 2010). Therefore, algorithms developed using datasets exclusively containing either children or adults may restrict their generalizability across diverse age ranges as the variability may be seen as image degradation (Vogelbacher et al., 2018).

### 4.4. Benchmark

To evaluate algorithm performance, a benchmark was established for all algorithms. Either visual QC or synthetically degraded images have been used for this goal. For visual QC, a predefined protocol is used to mitigate subjectivity (Backhausen et al., 2016), as there are inherent variations in determining an acceptable level of image quality (Scheltens et al., 1995). In case of synthetically degraded images, filters are applied to simulate various types of artifacts. For reproducibility purposes, the details of the filters used and the corresponding parameters employed should be specified. However, some studies lack information on their protocols, such as the number of raters and consensus methods. In none of the studies using synthetically degraded images, a rationale was provided for filter selection or parameter scaling.

To evaluate the visual QC output, most studies (Alfaro-Almagro et al., 2018; Fantini et al., 2021; Keshavan et al., 2019; Mortamet et al., 2009; Pizarro et al., 2016; Sujit et al., 2019; White et al., 2018) use a binary pass or fail classification, while two studies (Esteban et al., 2017; Kim et al., 2019) introduce a ’doubtful’ category, which is later merged with the pass category. This decision is based on achieving higher agreement between automatic QC and visual QC. However, it might be more advantageous to merge the ’doubtful’ category with the fail category, rather than the pass category, allowing users to focus their visual QC solely on the failed scans in case of semi-automatic QC (Alfaro-Almagro et al., 2018; Backhausen et al., 2016; Kim et al., 2019). Synthetically degraded images on the other hand, are measured on a continuous scale, often through percentages or scaling of the filter parameters. However, it remains unclear how this scaling relates to real-world artifacts, raising questions about the transferability and practical interpretation of these synthetic measures.

Both methods are limited in that there is a lack of consistency in the benchmark and thus direct comparisons and generalizations of the algorithms are hindered. In the case of the visual QC, there have been attempts to develop uniform and robust visual QC rating systems to improve replication and comparability between studies (Backhausen et al., 2016). In case of synthetically degraded images, there are concerns about the generalizability of the findings to real-world clinical settings, where the types and severity of artifacts may differ significantly from those artificially induced.

### 4.5. Future directions

To evaluate and compare algorithms in a non-biased manner, the algorithms should be trained on the same dataset. This way, it is ensured that comparisons between algorithms are focused on the inherent capabilities and limitations of the algorithms themselves rather than being influenced by specific characteristics of individual datasets. By encompassing a wide range of ages, pathologies, and artifact prevalence, the generalizability and performance across different scenarios might improve. Additionally, a consensus (Fantini et al., 2021) should be reached, for which the development of standardized protocols for visual QC or synthetically degraded images could be beneficial. Also, it might be useful to work with a benchmark composed of both real-world scans that are visually checked and synthetically degraded images, since this would reduce the subjectivity but also increase the generalizability of findings to real-world clinical settings.

Furthermore, researchers continue to explore new features to improve the algorithms, aiming for more robust and discriminative measures. However, it is equally important to investigate the discriminative power of the individual features that are already being used, particularly for specific artifacts. Additionally, explainable deep learning could be used to gain insight into the decision-making process of the deep learning based algorithms, and thus the image characteristics such algorithms focus on. Understanding which feature or set of features is most effective in capturing specific artifacts can provide valuable insights for further refinement or the algorithms.

Finally, generative models have recently attracted much attention in quality control or anomaly detection, due to its unique ability to generate new data when there is a lack of data that represents the anomalous behavior and the ability to apply representation learning (Sabuhi et al., 2021). This might also be useful for the QC of T1w MRI scans.

### 4.6. Conclusion

To the best of the authors’ knowledge, this is the first review on (semi-)automatic QC algorithms for T1w MRI scans. The detected algorithms employ diverse approaches and features, with an emphasis on entropy measures. However, comparing algorithm performance was challenging due to dataset-specific optimization, inflating results and hindering cross-dataset comparisons. Also variability and missing information in the benchmark was found, including unclearities in protocols and limited information on filter selection and parameter scaling. Despite these limitations, our review provides valuable insights into the landscape of QC algorithms for structural T1w scans. The implications of these findings call for future research and collaboration to establish guidelines and best practices, to ultimately enhance the reliability and effectiveness of QC algorithms in this domain.

## Data Availability

All data produced in the present work are contained in the manuscript

