## Supplemental Table 1 for "A systematic review of (semi-)automatic quality control of T1-weighted MRI scans"

Table S.1.: Search strategy in PubMed

| **Search** | **Query** | **Results** |
| --- | --- | --- |
| #7 | #5 NOT #6 | 268 |
| #6 | "Review"[Publication Type] OR "Systematic Review" [Publication Type] OR "Meta-Analysis"[Publication Type] OR "Meta-Analysis as Topic"[Mesh] OR "meta-analysis"[tiab] OR "systematic review*"[tiab] OR "systematic literature review*"[tiab] OR "Letter"[Publication Type] OR "Editorial"[Publication Type] OR "Comment"[Publication Type] | 5,402,824 |
| #5 | #1 AND #2 AND #3 AND #4 | 326 |
| #4 | "Neurosciences"[Mesh] OR "Brain"[Mesh] OR "brain"[tiab] OR "head"[tiab] OR "cereb*"[tiab] OR Neuro*[tiab] OR "grey matter"[tiab] OR "gray matter"[tiab] | 3,713,585 |
| #3 | "Magnetic Resonance Imaging"[Mesh] OR "magnetic resonance imag*"[tiab] OR "MRI"[tiab] OR "MR imag*"[tiab] OR "NMR"[tiab] OR "nuclear magnetic resonance"[tiab] OR Neuroimag*[tiab] OR "Neuro imag*"[tiab] | 953,913 |
| #2 | "Quality Control"[Mesh] OR "quality control*"[tiab] OR "quality assessment*"[tiab] OR "quality check*"[tiab] OR "quality evaluat*"[tiab] | 127,939 |
| #1 | "Automation"[Mesh] OR Automat*[tiab] OR Toolbox*[tiab] OR Software*[tiab] OR Program*[tiab] OR "Artificial Intelligence"[Mesh] OR "Bayes Theorem"[Mesh] OR "Markov Chains"[Mesh] OR "Latent Class Analysis"[Mesh] OR AdaBoost[tiab] OR AI[tiab] OR Artificial-Intelligence*[tiab] OR autoencoder*[tiab] OR auto-encoder*[tiab] OR Back-propagation*[tiab] OR Bayesian-learning[tiab] OR gradient-boosting[tiab] OR CART[tiab] OR classification-algorithm*[tiab] OR Computational-Intelligen*[tiab] OR Computer-heuristic*[tiab] OR Computer-reasoning*[tiab] OR Computer-vision*[tiab] OR Connectionist-model*[tiab] OR decision-stump*[tiab] OR deep-belief-network*[tiab] OR Deep-learning[tiab] OR dirichlet[tiab] OR elastic-net[tiab] OR Ensemble[tiab] OR Expert-system*[tiab] OR Fuzzy-system[tiab] OR gaussianprocess*[tiab] OR generalized-additive-model*[tiab] OR generative-adversarial-network*[tiab] OR genetic-algorithm*[tiab] OR gradient-response-unit*[tiab] OR GRU[tiab] OR Heuristic-learning[tiab] OR Hierarchical-learning*[tiab] OR hierarchical-temporal-memor*[tiab] OR Image-Interpretation*[tiab] OR Image-recognition[tiab] OR Kernel-method*[tiab] OR LARS[tiab] OR LASSO[tiab] OR latent-class*[tiab] OR latent-process*[tiab] OR latent-variable*[tiab] OR LDA[tiab] OR Learning-algorithm*[tiab] OR learning-automat*[tiab] OR learning-machine*[tiab] OR learning-vector-quanti*[tiab] OR Least-Absolute-Shrinkage-and-Selection-Operator[tiab] OR least-angle-regression*[tiab] OR Logitboost[tiab] OR Long-short-term-memory[tiab] OR LSTM[tiab] OR Machine-intelligen*[tiab] OR Machine-learning[tiab] OR Machine-vision*[tiab] OR Markov-model*[tiab] OR Naïve-Bayes[tiab] OR Neural-network*[tiab] OR Deep-Boltzmann-Machine*[tiab] OR partial-least-squares-regression*[tiab] OR penalized-regression*[tiab] OR perceptron[tiab] OR PLSregression*[tiab] OR QDA[tiab] OR Qlearning[tiab] OR Q-learning[tiab] OR quadratic-classifier*[tiab] OR quadratic-discriminant*[tiab] OR random-forest*[tiab] OR reinforcement-learning[tiab] OR ridge-regression*[tiab] OR Rule-based[tiab] OR self-organising-map*[tiab] OR Speech-recognition[tiab] OR stacked-generali*[tiab] OR Supervised-learning[tiab] OR Support-vector-machine*[tiab] OR Support-vector-machine*[tiab] OR temporal-difference-learning[tiab] OR Textmining[tiab] OR Text-mining[tiab] OR Transfer-learning[tiab] OR Unsupervised-learning[tiab] OR XAI[tiab] OR "Machine Learning"[Mesh] OR "Machine Learning"[tiab] OR "machine intelligen*"[tiab] OR "machine vision*"[tiab] OR "machine learning"[tiab] OR "transfer learning"[tiab] OR "deep learning"[tiab] OR "neural network*"[tiab] OR "support vector machine*"[tiab] OR "Long short term memory"[tiab] OR "LSTM"[tiab] OR "supervised learning"[tiab] OR "unsupervised learning"[tiab] OR "reinforcement learning*"[tiab] OR "hierarchical learning*"[tiab] OR "Prediction model*"[tiab] OR "perceptron"[tiab] | 1,945,046 |
