## Supplemental Table 2 for "A systematic review of (semi-)automatic quality control of T1-weighted MRI scans"

Table S.2.: Search strategy in Embase.com

| **Search** | **Query** | **Results** |
| --- | --- | --- |
| #6 | #5 AND ('article' / it OR 'article in press' / it OR 'conference paper' / it) | 496 |
| #5 | #1 AND #2 AND #3 AND #4 | 917 |
| #4 | 'neuroscience' / exp OR 'brain' / exp OR ('brain' OR 'head' OR 'cereb*' OR 'neuro*' OR 'grey matter' OR 'gray matter'):ti,ab,kw | 4,847,707 |
| #3 | 'nuclear magnetic resonance imaging' / exp OR ('magnetic resonance imag*' OR 'mri' OR 'mr imag*' OR 'nmr' OR 'nuclear magnetic resonance' OR 'neuroimag*' OR 'neuro imag*'):ti,ab,kw | 1,531,137 |
| #2 | 'quality control' / de OR (‘quality control*’ OR ‘quality assessment*’ OR ‘quality check*’ OR ‘quality evaluat*’):ti,ab,kw | 285,715 |
| #1 | 'automation' / exp OR 'automation':ti,ab,kw OR automat*:ti,ab,kw OR toolbox*:ti,ab,kw OR software*:ti,ab,kw OR program*:ti,ab,kw OR 'artificial intelligence' / exp OR 'artificial intelligence':ti,ab,kw OR 'bayes theorem' / exp OR 'bayes theorem':ti,ab,kw OR 'Markov chain' / exp OR 'markov chain*':ti,ab,kw OR 'latent structure analysis' / exp OR 'latent class analysis':ti,ab,kw OR adaboost:ti,ab,kw OR ai:ti,ab,kw OR 'artificial intelligence*':ti,ab,kw OR autoencoder*:ti,ab,kw OR 'auto encoder*':ti,ab,kw OR 'back propagation*':ti,ab,kw OR 'bayesian learning':ti,ab,kw OR 'gradient boosting':ti,ab,kw OR cart:ti,ab,kw OR 'classification algorithm*':ti,ab,kw OR 'computational intelligen*':ti,ab,kw OR 'computer heuristic*':ti,ab,kw OR 'computer reasoning*':ti,ab,kw OR 'computer vision*':ti,ab,kw OR 'connectionist model*':ti,ab,kw OR 'decision stump*':ti,ab,kw OR 'deep belief network*':ti,ab,kw OR dirichlet:ti,ab,kw OR 'elastic net':ti,ab,kw OR ensemble:ti,ab,kw OR 'expert system*':ti,ab,kw OR 'fuzzy system':ti,ab,kw OR gaussianprocess*:ti,ab,kw OR 'generalized additive model*':ti,ab,kw OR 'generative adversarial network*':ti,ab,kw OR 'genetic algorithm*':ti,ab,kw OR 'gradient response unit*':ti,ab,kw OR gru:ti,ab,kw OR 'heuristic learning':ti,ab,kw OR 'hierarchical temporal memor*':ti,ab,kw OR 'image interpretation*':ti,ab,kw OR 'image recognition':ti,ab,kw OR 'kernel method*':ti,ab,kw OR lars:ti,ab,kw OR lasso:ti,ab,kw OR 'latent class*':ti,ab,kw OR 'latent process*':ti,ab,kw OR 'latent variable*':ti,ab,kw OR lda:ti,ab,kw OR 'learning algorithm*':ti,ab,kw OR 'learning automat*':ti,ab,kw OR 'learning machine*':ti,ab,kw OR 'learning vector quanti*':ti,ab,kw OR 'least absolute shrinkage and selection operator':ti,ab,kw OR 'least angle regression*':ti,ab,kw OR logitboost:ti,ab,kw OR lstm:ti,ab,kw OR 'markov model*':ti,ab,kw OR 'naïve bayes':ti,ab,kw OR 'deep boltzmann machine*':ti,ab,kw OR 'partial least squares regression*':ti,ab,kw OR 'penalized regression*':ti,ab,kw OR perceptron:ti,ab,kw OR plsregression*:ti,ab,kw OR qda:ti,ab,kw OR qlearning:ti,ab,kw OR 'q learning':ti,ab,kw OR 'quadratic classifier*':ti,ab,kw OR 'quadratic discriminant*':ti,ab,kw OR 'random forest*':ti,ab,kw OR 'reinforcement learning':ti,ab,kw OR 'ridge regression*':ti,ab,kw OR 'rule based':ti,ab,kw OR 'self organising map*':ti,ab,kw OR 'speech recognition':ti,ab,kw OR 'stacked generali*':ti,ab,kw OR 'temporal difference learning':ti,ab,kw OR textmining:ti,ab,kw OR 'text mining':ti,ab,kw OR xai:ti,ab,kw OR 'machine learning' / exp OR 'machine learning':ti,ab,kw OR 'machine intelligen*':ti,ab,kw OR 'machine vision*':ti,ab,kw OR 'machine learning':ti,ab,kw OR 'transfer learning':ti,ab,kw OR 'deep learning':ti,ab,kw OR 'neural network*':ti,ab,kw OR 'support vector machine*':ti,ab,kw OR 'long short term memory':ti,ab,kw OR 'lstm':ti,ab,kw OR 'supervised learning':ti,ab,kw OR 'unsupervised learning':ti,ab,kw OR 'reinforcement learning*':ti,ab,kw OR 'hierarchical learning*':ti,ab,kw OR 'prediction model*':ti,ab,kw OR 'perceptron':ti,ab,kw | 2,706,198 |
