## Supplemental Table 3 for "A systematic review of (semi-)automatic quality control of T1-weighted MRI scans"

Table S.3.: Search strategy in Web of Science

| **Search** | **Query** | **Results** |
| --- | --- | --- |
| #6 | #4 AND #3 AND #2 AND #1 and Article or Early Access or Data Paper (Document Types) | 252 |
| #5 | #1 AND #2 AND #3 AND #4 | 298 |
| #4 | TS=("brain" OR "head" OR "cereb*" OR Neuro* OR "grey matter" OR "gray matter") | 4,012,639 |
| #3 | TS=("magnetic resonance imag*" OR "MRI" OR "MR imag*" OR "NMR" OR "nuclear magnetic resonance" OR Neuroimag* OR "Neuro imag*") | 1,208,383 |
| #2 | TS=( "quality control*" OR "quality assessment*" OR "quality check*" OR "quality evaluat*") | 161,548 |
| #1 | TS=(Automat* OR Toolbox* OR Software* OR Program* OR AdaBoost OR AI OR Artificial-Intelligence* OR autoencoder* OR auto-encoder* OR Back-propagation* OR Bayesian-learning OR gradient-boosting OR CART OR classification-algorithm* OR Computational-Intelligen* OR Computer-heuristic* OR Computer-reasoning* OR Computer-vision* OR Connectionist-model* OR decision-stump* OR deep-belief-network* OR Deep-learning OR dirichlet OR elastic-net OR Ensemble OR Expert-system* OR Fuzzy-system OR gaussianprocess* OR generalized-additive-model* OR generative-adversarial-network* OR genetic-algorithm* OR gradient-response-unit* OR GRU OR Heuristic-learning OR Hierarchical-learning* OR hierarchical-temporal-memor* OR Image-Interpretation* OR Image-recognition OR Kernel-method* OR LARS OR LASSO OR latent-class* OR latent-process* OR latent-variable* OR LDA OR Learning-algorithm* OR learning-automat* OR learning-machine* OR learning-vector-quanti* OR Least-Absolute-Shrinkage-and-Selection-Operator OR least-angle-regression* OR Logitboost OR Long-short-term-memory OR LSTM OR Machine-intelligen* OR Machine-learning OR Machine-vision* OR Markov-model* OR Naïve-Bayes OR Neural-network* OR Deep-Boltzmann-Machine* OR partial-least-squares-regression* OR penalized-regression* OR perceptron OR PLSregression* OR QDA OR Qlearning OR Q-learning OR quadratic-classifier* OR quadratic-discriminant* OR random-forest* OR reinforcement-learning OR ridge-regression* OR Rule-based OR self-organising-map* OR Speech-recognition OR stacked-generali* OR Supervised-learning OR Support-vector-machine* OR Support-vector-machine* OR temporal-difference-learning OR Textmining OR Text-mining OR Transfer-learning OR Unsupervised-learning OR XAI OR "Machine Learning" OR "machine intelligen*" OR "machine vision*" OR "machine learning" OR "transfer learning" OR "deep learning" OR "neural network*" OR "support vector machine*" OR "Long short term memory" OR "LSTM" OR "supervised learning" OR "unsupervised learning" OR "reinforcement learning*" OR "hierarchical learning*" OR "Prediction model*" OR "perceptron") | 4,161,582 |
